# Real-world clinical performance of commercial SARS-CoV-2 rapid antigen tests in suspected COVID-19: A systematic meta-analysis of available data as per November 20, 2020

**DOI:** 10.1101/2020.12.22.20248614

**Authors:** Johannes Hayer, Dusanka Kasapic, Claudia Zemmrich

## Abstract

**Background:** Immunochromatographic rapid antigen tests (RATs) emerged onto the COVID-19 pandemic testing landscape to aid in the rapid diagnosis of people with suspected SARS-CoV-2 infection. RATs are particularly useful where RT-PCR is not immediately available and symptoms suggestive of a high viral load and infectiousness are assumed. Several lateral flow immunoassays have been authorized for use under EUA and/or the CE mark, presenting varying overall clinical performance data generated by the manufacturer or by independent investigators. To compare the real-world clinical performance of commercially available rapid chromatographic immunoassays intended for the qualitative detection of SARS-CoV-2, we performed a systematic meta-analysis of published data.

**Methods:** We searched the MEDLINE®, Embase, BIOSIS and Derwent Drug File (ProQuest)for manufacturer-independent prospective clinical performance studies comparing SARS-CoV-2 RATs and RT-PCR assays. Only studies on lateral flow assays not needing a separate reader for retrieving the result were included, if data were available on viral load, patients’ symptom status, sample type, and PCR assay used. For better data comparability, recalculation of the studies’ single performance data confidence intervals using the exact Clopper–Pearson method was applied.

**Results:** We could include 19 studies (ten peer-reviewed) presenting detailed clinical performance data on 11,209 samples with 2449 RT-PCR-positives out of study prevalence rates between 1.9–100 % and between 50– 100% symptomatic samples. Four studies directly compared two to three different RATs and 15 studies compared one RAT to RT-PCR. Overall specificity ranged, with one test outlier, between 92.4% (87.4– 95.9) and 100% (99.7–100), and overall clinical sensitivity varied between 28.9% (16.4–44.3) and 98.3% (91.1–99.7), depending on assay, population characteristics, viral load, and symptom status. Sensitivity in high-viral-load samples (cycle threshold ≤25) showed a considerable heterogeneity among the assays ranging from 66.7% to 100%.

**Conclusion:** Only two RATs offered sufficient manufacturer-independent, real-world performance data supporting use for the detection of current SARS-CoV-2 infection in symptomatic or high-viral-load patient populations. Reliable positive predictive values require testing of symptomatic patients or asymptomatic individuals only in case of a high pre-test probability. If RATs are used for screening of asymptomatic cases in low-prevalence scenarios, a lower positive predictive value of the result has to be considered.

## Background

Real-time reverse transcriptase PCR (RT-PCR) is the current gold standard to detect acute SARS-CoV-2 infection using nasopharyngeal, nasal, or oropharyngeal swab samples containing a measurable amount of viral RNA. It requires a professionally run laboratory with molecular-biological competence, as well as transport infrastructure between the place of sample collection and the lab. Immunochromatographic rapid antigen tests (RATs) emerged on the pandemic testing landscape to support the rapid diagnosis of individuals suspected of SARS-CoV-2 infection, either by presenting symptoms or by contact with cases. These point-of-care tests are less clinically sensitive than RT-PCR tests but overall offer a reliable specificity. Several lateral flow immunoassays have been authorized for use under EUA and/or the CE mark,^1,2^ presenting clinical performance data generated mostly by the manufacturer and containing heterogeneous patient populations. The Foundation for Innovative New Diagnostics (FIND) collates new diagnostic SARS-CoV-2 test developments and manufacturer-independent validation studies of newly commercialized tests on a regularly updated webpage.^3^

Sensitivity and specificity of RATs depend on numerous variables contributing to the final test results. Their lower limit of detection is at a higher viral load compared with RT-PCR tests.^4^ Most RATs are intended for use in patients up to 5 to 7 days after symptom onset, to increase the probability of having sufficiently high viral load for detection. If RATs are used to assess asymptomatic contact samples, time from symptom onset is not available and can only be assumed by the date of the index case contact. The use of a RAT for screening within a low-prevalence population may not be appropriate, as fewer cases with a detectable high viral load are expected within this group, decreasing the positive predictive value of the test accordingly.^5^

Direct assay comparison studies minimize further heterogeneity parameters (e.g. PCR assay performance differences or varying circulating virus cell characteristics) but are limited due to the invasive character of repeat sample extraction and a high number of required screened persons to detect a sufficient number of positive cases.

In order to provide clarity on the real-world clinical performance of RATs, we compiled all available manufacturer-independent, prospectively collected, clinical data using rapid chromatographic immunoassays, which were commercially available as of November 20, 2020 and intended for the qualitative detection of SARS-CoV-2 present in human nasopharynx, in individuals suspected of SARS-CoV-2 infection. We aimed to harmonize the data regarding the aforementioned performance-impacting factors as much as possible using mathematical methods, to ensure that the data are to a minimum comparable, despite varying methods of presentation in the publications considered for this analysis.

## Materials and Methods

### Search strategy

We searched MEDLINE®, EMBASE, BIOSIS, and Derwent Drug File at Host “ProQuest” for any clinical performance studies using a commercial SARS-CoV-2 RAT for the following search terms: “COVID-19” OR “SARS-CoV-2” OR “2019-nCoV” OR “coronavirus disease 2019” OR “novel corona virus” OR MESH Entries for Coronaviridae (incl. narrow terms) OR EMTREE Entries for Coronaviridae (incl. narrow terms) OR MESH/EMTREE Entries for “severe acute respiratory syndrome” (incl. narrow terms) AND “rapid antigen test*” OR “rapid antigen assay*” OR “standard Q covid-19 ag” AND “sensitivity” OR “specificity” OR “clinical performance” OR “positive agreement” OR “negative agreement” OR “concordance” OR “validation” OR “evaluation” OR “accuracy”. After exclusion of 40 findings not representing clinical trials (29 findings) or not meeting our inclusion criteria (11 findings), eight clinical trials were extracted, along with two further peer-reviewed trial manuscripts that were found manually. Secondly, we searched the studies listed on the Foundation for Innovative Diagnostics (FIND) website (only final, no interim study reports were considered), the European Commission (EC) COVID-19 Diagnostic Devices and Test Method database website (both accessed December 10, 2020), and the diagnosticsglobalhealth.org website (accessed December 14, 2020); nine pre-print clinical RAT studies were added to our finding list. A summary of the search result is presented in Figure 1.

**Figure 1.**
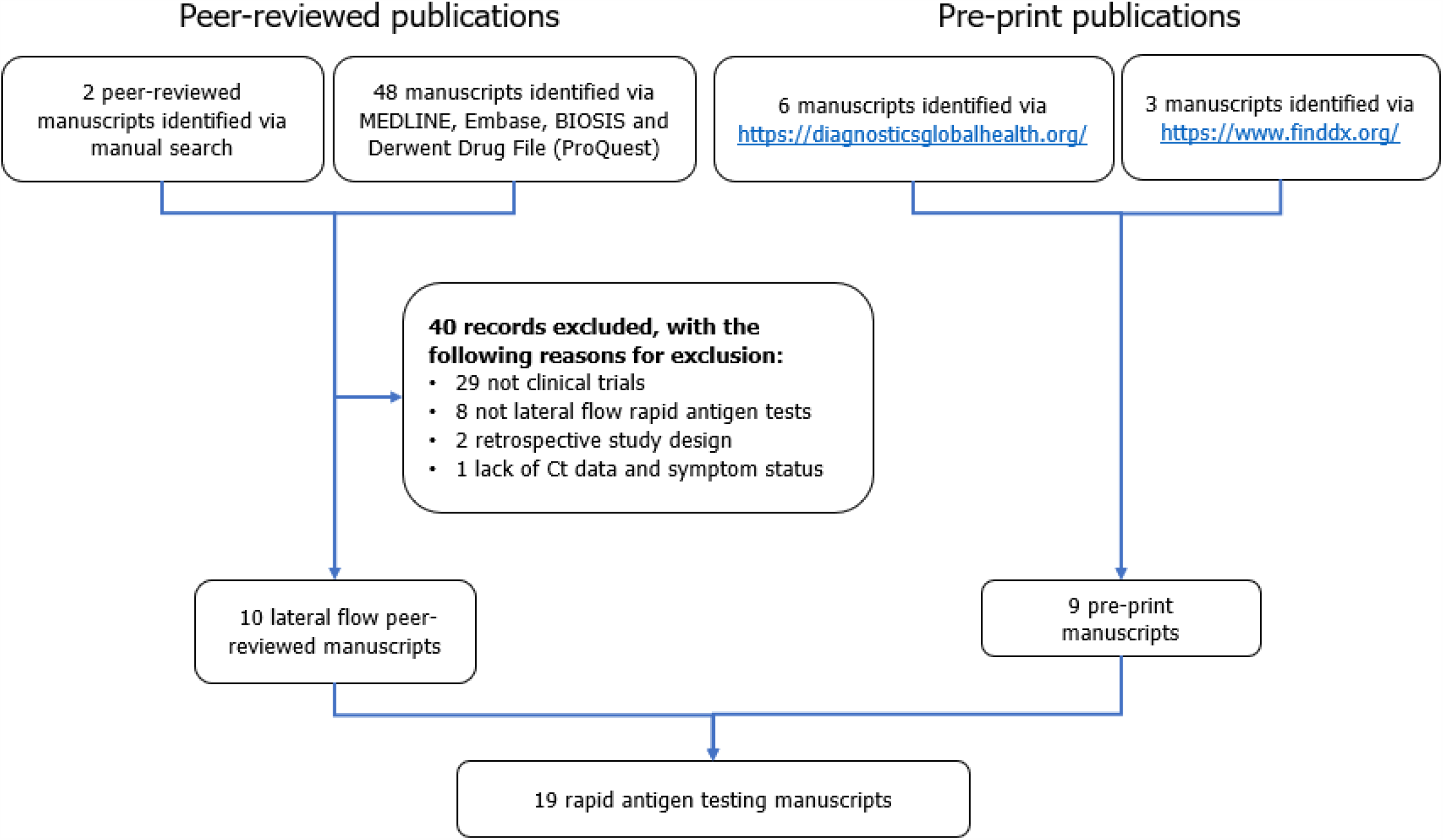
Flowchart of search results.

### Selection criteria

Only studies investigating clinical performance data of RATs not requiring a separate reading device compared to RT-PCR were considered, if they were performed independent of funding by the manufacturer or distributor. Retrospective laboratory studies were excluded, information on cycle threshold (Ct) values and symptom status had to be available. To ensure comparability, only nasopharyngeal or combined oro- and nasopharyngeal sample types were considered, either directly performed at the point-of-care or at a laboratory after sample transport in viral transport media. If stated, the target gene(s) of the RT-PCR assay was/were reported. No further exclusion criteria on the studies considered for this analysis have been defined. Parameters such as different classification of a negative sample result according to Ct >38 or >40, or the definition of symptoms qualifying as a “symptomatic patient suspected of COVID-19” varied between the studies.

The data were extracted to an electronic database and stratified according to the testing devices. Clinical performance data per Ct group, and stratification for symptoms versus no symptoms present, were calculated from the single data in case they were available. Finally ten peer-reviewed studies and nine pre-print manuscripts could have been included in the meta-analysis, investigating five different RATs: the SARS-CoV-2 Rapid Antigen Test (Roche), equaling the STANDARD Q COVID-19 Ag Test (SD Biosensor, henceforth called “Roche/SDB”); the Panbio™ COVID-19 Ag Test (Abbott, henceforth called “Abbott”); the COVID-19 Ag Respi-Strip® (Coris Bioconcept, henceforth called “Coris”); the COVID-Viro® (AAZ-LMB, henceforth called “AAZ”); and the BIOCREDIT COVID-19 Ag (RapiGEN, henceforth called “RapiGEN”).

### Statistical analyses

Performance results from the comparison of the RATs against the RT-PCR methods are reported as sensitivity and specificity. If available, sensitivity is also provided for cohorts dependent on the Ct values. As confidence intervals reported in the manuscripts used different methods, all confidence intervals have been recalculated for our review using the exact Clopper–Pearson method to allow for better comparability.

The meta-analysis of the performance results of the RATs against the RT-PCR methods was performed for the Roche/SDB, Abbott, and Coris RATs using the statistical software R.

The metaprop function from the “meta” package was used to calculate the effect size for each different test and overall. The results of the AAZ and RapiGEN RATs are also included, even though only one study was available for those tests. The results are shown as a forest plot summarizing the sensitivities found in the different studies.

The bivariate model of Reitsma et al.^6^ was fitted as a linear mixed model and variance components are estimated by restricted maximum likelihood (REML), using the reitsma function from the “mada” package for each system investigated in more than one study. The results are presented as a summary receiver operating characteristic (SROC) curve plot^7^ including the results of all systems (also those investigated in only one study). The single studies, summary estimates, SROC curves, and confidence regions are depicted.

The relationship between sensitivity and viral load represented by Ct value is visualized in a scatterplot. The single study results for sensitivity below a certain Ct-threshold are plotted against these Ct values categorized by the different systems. If in a single study sensitivity estimates for more than one Ct value were available, those are connected by a line.

## Results

### Study population characteristics

Altogether, we included data from 19 clinical studies providing single data on 11,209 samples, including 2449 samples with confirmed SARS-Cov-2 by RT-PCR, see Tables 1–4. Three studies directly compared the Roche/SDB RAT with the Abbott,^8^ Coris,^9^ or RapiGEN^10^ RATs, respectively, and one study compared the Roche/SDB RAT with both the Abbott and the AAZ RAT,^11^ all against RT-PCR. 15 studies compared the clinical performance of one RAT against RT-PCR: six studies compared the Roche/SDB;^12-17^ six compared the Abbott;^18-23^ and three compared the Coris RAT.^24-26^ All studies provided descriptions of the study populations, regarding mean age and gender distribution (data not shown), as well as symptoms, prevalence rates, and Ct of the RT-PCR-positive samples. Prevalence rate – here meaning the number of RT-PCR-positive samples within the overall study population – varied between 1.9–100%. The prevalence of SARS-CoV-2 in some of these studies did not reflect the prevalence in the local populations, as additional pre-specified testing criteria qualified patients for study entry and created a preselection bias.

**Table 1.** Study population characteristics and overall performance: Roche/SDB only.

|  | Chaimayo<sup>12</sup> | Igloi<sup>15</sup> | Lindner<sup>16</sup> | Cerutti<sup>13</sup> | Krüttgen<sup>14</sup> | Nalumansi<sup>17</sup> |
| --- | --- | --- | --- | --- | --- | --- |
| **Country** | Thailand | Netherlands | Germany | Italy | Germany | Uganda |
| **Participants (N)** | 454 | 970 | 289 | 330 | 150 | 262 |
| **PCR+ (n)** | 60 | 186 | 39 | 109 | 75 | 90 |
| **Prevalence (%)** | 13.2 | 19.2 | 13.5 | 33 | 50 | 34.4 |
| **Symptomatic (%)** | 95.0 (PCR+) | 91.3 (overall) | 97.6 (overall) | 56.1 (overall);<br>95.4 (PCR+) | Not reported | 14 (PCR+) |
| **PCR manufacturer and assay** | Seegene Allplex™ 2019-nCoV Assay | Roche cobas® SARS-CoV-2 Assay | Roche cobas® SARS-CoV-2 Assay;<br>TIB MOLBIOL SARS-CoV-2 E-gene® | Roche cobas® SARS-CoV-2 Assay;<br>Seegene Allplex™ 2019-nCoV Assay;<br>DiaSorin Simplexa® | Altona Diagnostics RealStar® SARS-CoV-2 | Metabion custom oligo nucleotides |
| **Specimen Ct values (min – max); Mean/Median** | 10.5–39.0;<br>Mean 22.8±6.7 (E-gene) | 15.6–37.4;<br>Median 23.6 | 17.3–35.5;<br>Mean 23.7±5.5;<br>Median 21.9 | 12.3–38.1 | <20–≥ 35 | ≤29–39 |
| **Antigen test performance** |  |  |  |  |  |  |
| **Overall specificity (95% CI)<sup>#</sup>** | 98.7 (97.1–99.5) | 99.5 (98.7–99.9) | 99.6 (97.8–100) | 100 (98.3–100) | 96.0 (88.8–99.2) | 92.4 (87.4–95.9) |
| **Overall sensitivity (95% CI)<sup>#</sup>** | 98.3 (91.1–99.7) | 84.9 (79.0–89.8) | 79.5 (63.5–90.7) | 70.6 (61.2–79.0) | 70.7 (59.0–80.6) | 70 (59.4–79.2) |
| **Sensitivity by Ct (95% CI)<sup>#</sup>** | Ct≤31: 100 | Ct≤25: 99.1 (95.2–100);<br>Ct≤30: 94.3 (89.6–97.0) | Ct≤24: 100 (85.2–100);<br>Ct≤25.3: 96.2 (80.4–99.9);<br>Ct≤29.6: 90.3 (74.2–98.0);<br>Ct≤32: 88.6 (73.3–96.8) | Ct<28: 100;<br>Ct 28–30: 38.5;<br>Ct 30–35: 26.7;<br>Ct>35: 9.1 | Ct<25:100;<br>Ct 25–<30: 95.0;<br>Ct 30–<35: 44.8;<br>Ct>=35: 22.2 | Ct≤29: 91.9% (78.1–98.3);<br>Ct 30–37: 54.5%;<br>Ct 38–39: 55.6% |

<sup>#</sup>Values were recalculated using the original data and confidence intervals were calculated using the exact Clopper–Pearson method.

**Table 2.**
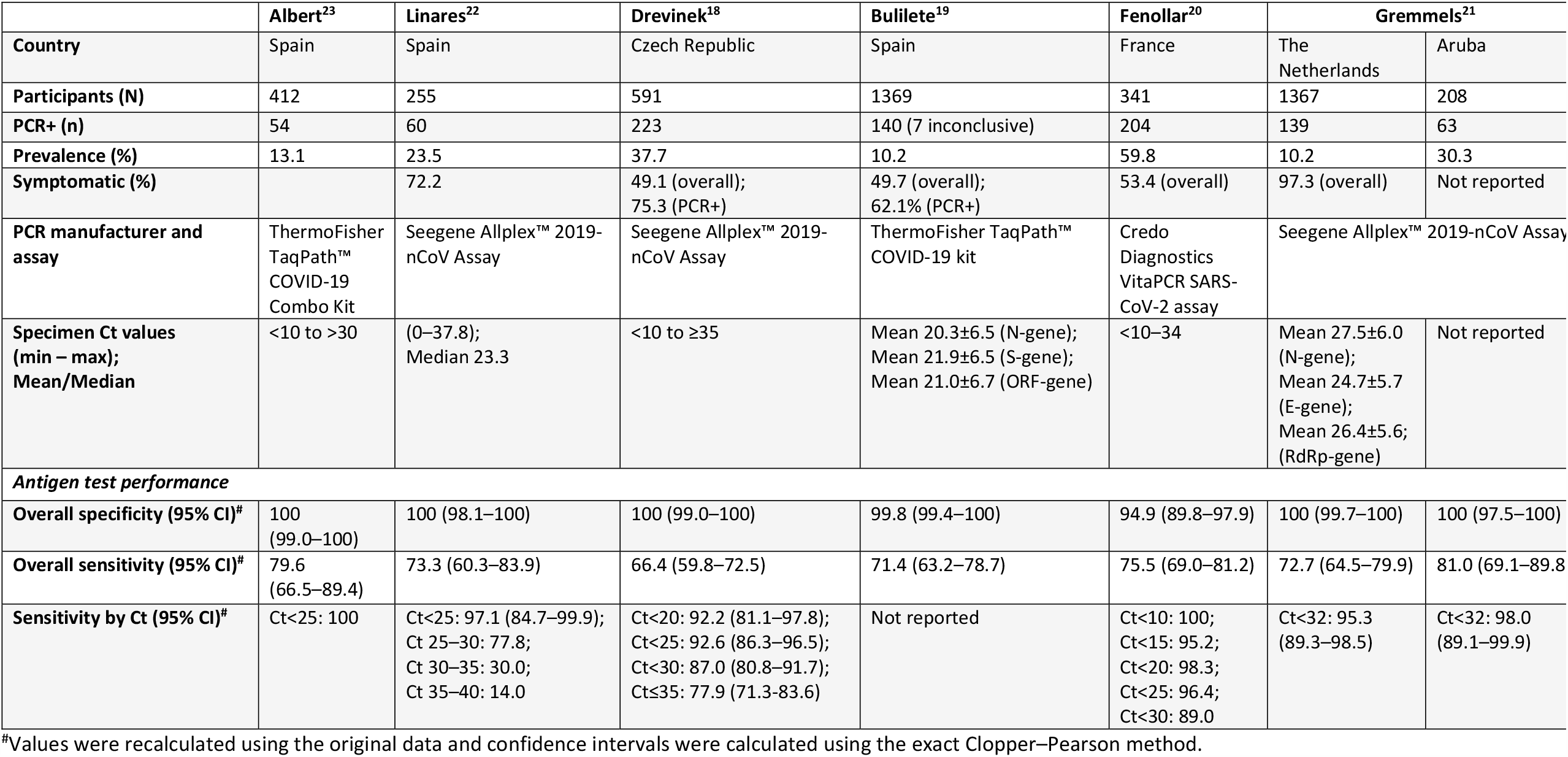
Study population characteristics and overall performance – Panbio, Abbott only.

**Table 3.** Study population characteristics and overall performance – CORIS RESPI-strip only.

|  | Lambert-Niclot<sup>24</sup> | Veyrenche<sup>25</sup> | Schohy<sup>26</sup> |
| --- | --- | --- | --- |
| **Country** | France | France | Belgium |
| **Participants (N)** | 138 | 45 | 148 |
| **PCR+ (n)** | 94 | 45 | 106 |
| **Prevalence (%)** | 68.1 | NA (only positive clinical samples) | 71.6 |
| **Symptomatic (%)** | Not reported | 100 | 88.5 (overall) |
| **PCR manufacturer and assay** | Altona Diagnostics RealStar® SARS-CoV-2; Anatolia Geneworks Bosphore novel coronavirus (2019 nCoV) detection kit; Roche cobas® SARS-CoV-2 Assay; Seegene Allplex™ 2019-nCoV Assay | Seegene Allplex™ 2019-nCoV Assay | Primerdesign genesig® Real-Time PCR coronavirus COVID-19 |
| **Specimen Ct values (min – max); Mean/Median** | ≤10 to >40 | <15 to >40 | (16–38); Mean 31.4; Median 33 |
| **Antigen test performance** |  |  |  |
| **Overall specificity (95% CI)<sup>#</sup>** | 100 (92.0–100) | NA | 100 (91.6–100) |
| **Overall sensitivity (95% CI)<sup>#</sup>** | 50.0 (39.5–60.5) | 28.9 (16.4–44.3) | 30.2 (21.7–39.9) |
| **Sensitivity by Ct (95% CI)<sup>#</sup>** | Ct<25: 82.2 | Ct≤25: 86.7 (59.5–98.3);<br>Ct>25: 0.0 (0.0–11.6) | Ct<25: 100;<br>Ct<30: 70.6;<br>Ct<35: 46.9 |

<sup>#</sup>Values were recalculated using the original data and confidence intervals were calculated using the exact Clopper–Pearson method.

**Table 4.** Study population characteristics and overall performance – studies assessing multiple antigen assays.

|  | Schwob<sup>11</sup> |  |  | Berger<sup>8</sup> |  | Krüger<sup>9</sup> |  | Khairat<sup>10</sup> |  |
| --- | --- | --- | --- | --- | --- | --- | --- | --- | --- |
| Country | Switzerland |  |  | Switzerland |  | Germany + UK |  | Egypt |  |
| Antigen assay | Roche/SDB | Panbio, Abbott | COVID-Viro AAZ-LMB | Panbio, Abbott | Roche/SDB | Roche/SDB | CORIS RESPI-strip | Roche/SDB | BioCREDIT |
| Participants (N) | 928 |  |  | 1064 |  | 1688* |  | 100 |  |
| Participants per assay (n) | 333 | 271 | 324 | 535 | 529 | 1263 | 425 | 100 | 100 |
| PCR+ (n) | 112 | 122 | 138 | 124 | 191 | 47 | 8 | 80 | 80 |
| Prevalence (%) | 33.6 | 45.0 | 42.6 | 23.2 | 36.1 | 3.7 | 1.9 | 75 | 75 |
| Symptomatic (%) | 96 (overall) |  |  | 97.8 (PCR+) |  | 84.4 (overall) | 68.9 (overall) | Unreported |  |
| PCR manufacturer and assay | Roche cobas® SARS-CoV-2 Assay; in-house RT-PCR |  |  | Roche cobas® SARS-CoV-2 Assay |  | Germany: TIB MOLBIOL SARS-CoV-2 E-gene; Abbott RealTime SARS-CoV-2 assay; Roche cobas® SARS-CoV-2 Assay; Seegene Allplex™ 2019-nCoV Assay<br>UK: Primerdesign genesig® Real-Time PCR coronavirus COVID-19 |  | Unreported |  |
| Specimen Ct values (min – max); Mean/Median | NA<br>(Results per viral load available) |  |  | (14.2–39.7); Mean 22.5±5.1; Median 21.5 |  | NA | NA | Median 18.57 |  |
|  |  |  |  | (14.2–39.7); Mean 22.4±5.4; Median 21.8 | (14.4–37.4); Mean 22.6±4.9; Median 21.0 |  |  |  |  |
| Antigen test performance |  |  |  |  |  |  |  |  |  |
| Overall specificity (95% CI)<sup>#</sup> | 100 (98.3–100) | 100 (97.6–100) | 100 (98.0–100) | 100 (99.1–100) | 99.7 (98.3–99.9) | 99.3 (98.6–99.7) | 95.8 (93.4–97.6) | 95 (75.1–99.9) | 45 (23.1–68.5) |
| Overall sensitivity (95% CI)<sup>#</sup> | 92.9 (86.4–96.9) | 86.1 (78.6–91.7) | 84.1 (76.9–89.7) | 85.5 (78.0–91.2) | 89.0 (83.7–93.1) | 76.6 (62.0–87.7) | 50.0 (21.5–78.5) | 68.8 (57.4–78.7) | 52.5 (41.0–63.8) |
| **Sensitivity by Ct (95% CI)<sup>#</sup>** |  |  |  |  |  | Ct<25: 100 (81.5–100);<br>Ct≥25: 62.1 (42.3–79.3) | Ct<25:66.7 (9.4–99.2);<br>Ct≥25:40.0 (5.3–85.3) | Ct<18.57: 77.5 (61.5–89.2);<br>Ct>18.57: 60.0 (43.3–75.1) | Ct<18.57: 60.0 (43.3–75.1);<br>Ct>18.57: 45.0 (29.3–61.5) |

\*Total sample number of study was 2417, but one assay was excluded from our analysis for not fulfilling inclusion criteria; <sup>#</sup>Values were recalculated using the original data and confidence intervals were calculated using the exact Clopper–Pearson method;

Sample characterization according to Ct value of the RT-PCR-positive samples showed a mean Ct value between 20–33. The definition of high viral load varied considerably within the trials, from Ct <18.577 in an Egyptian study^10^ and <37 in a Ugandan trial.^17^ The Ct groups, which were summarized and analyzed by group, also differed considerably. A majority, but not all, have chosen to report groups of Ct ≤20, ≤25, ≤30, and ≤35. The cobas® (Roche) and Allplex / Seegene® were the most frequently used RT-PCR assays, but even within a single study up to five different assays were used.^9^ Mostly the E-gene was detected, sometimes the RdRp-gene or N-gene as well, and sometimes the target was not stated. One study did not report Ct values but reported RNA copies/mL.^11^

Local definitions of “patients suspected of SARS-CoV-2 infection” either included only patients presenting clinical symptoms, or also asymptomatic persons with recent direct contact with suspected or confirmed cases. Some asymptomatic contact case groups were limited to healthcare professionals with recent contact, but others included travelers returning from risk areas, which meant that this population varied in terms of pretest probability. The proportion of symptomatic versus asymptomatic samples highly varied, ranging between 14.0–97.8%. Not every report clearly stated the ratio between symptomatic RT-PCR-positive and symptomatic RT-PCR-negative samples. This ratio seemed to differ considerably; some studies reported overall >90% symptomatic persons gaining approximately 15% RT-PCR-positive samples, and the study which investigated only 14% symptomatic persons tested 34.4% RT-PCR-positive samples.

### Clinical performance of the RATs

Overall specificity of the investigated RATs ranged between 92.4–100% with one outlier (45%), ^10^ see Tables 1–4. The two rapid test assays with the most comprehensive available database of >6 studies, the Roche/SDB and Abbott, reported a specificity of >97% in the majority of the according trials, meeting the requirement threshold defined by health authorities.^5,27^ The Coris RAT ranged between 95.8–100% for specificity but this was combined with unacceptable low sensitivity. The AAZ showed very good results with specificity of 100% and sensitivity of 84.1% but was only evaluated in a single study and RapiGEN showed unacceptable low specificity of 45% in the only published study, combined with low sensitivity as well.

Pooled reported overall sensitivities ranged between 37.7–84.1% (95% CI 27.8–48.7 and 77.0–89.3 respectively), demonstrating considerable heterogeneity between the assays but better comparability within the assays, see Figure 2. Only the AAZ, Roche/SDB and Abbott tests showed a comparable overall performance and comply with Health Authorities’ requirement of a sensitivity >80%,^5,27^ see according SROC plots in Figure 3. Expectedly, all assays performed better in samples with high viral loads, but sensitivity dropped more rapidly at Ct >20 for the Coris and RapiGEN tests and less rapidly for the Roche/SDB and Abbott tests, see Figure 4. All assays showed a low sensitivity at Ct >30–32 and variable accuracy at Ct between 25–30.

**Figure 2.**
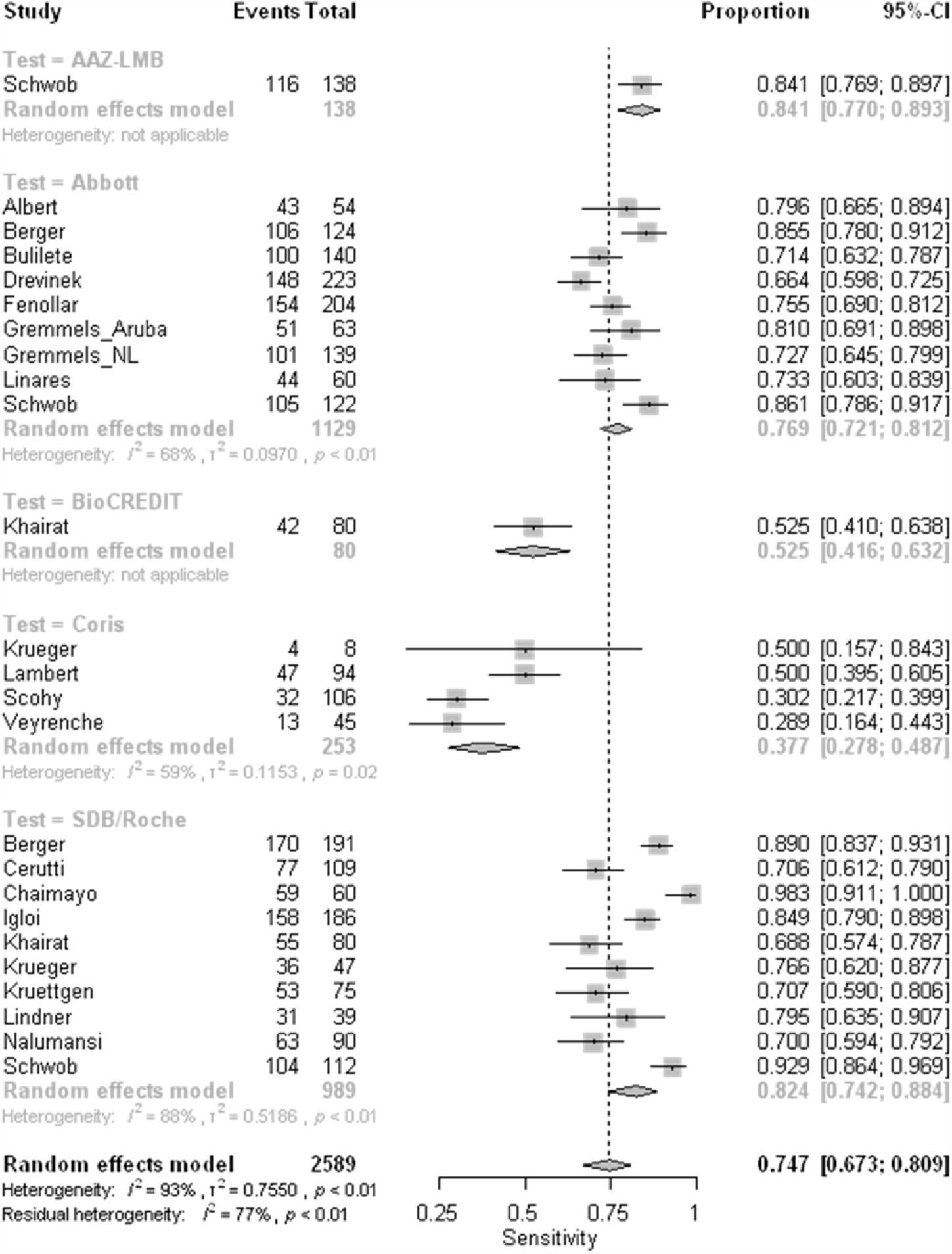
Forest plot of studies evaluating rapid antigen test sensitivity, grouped by test.

**Figure 3.**
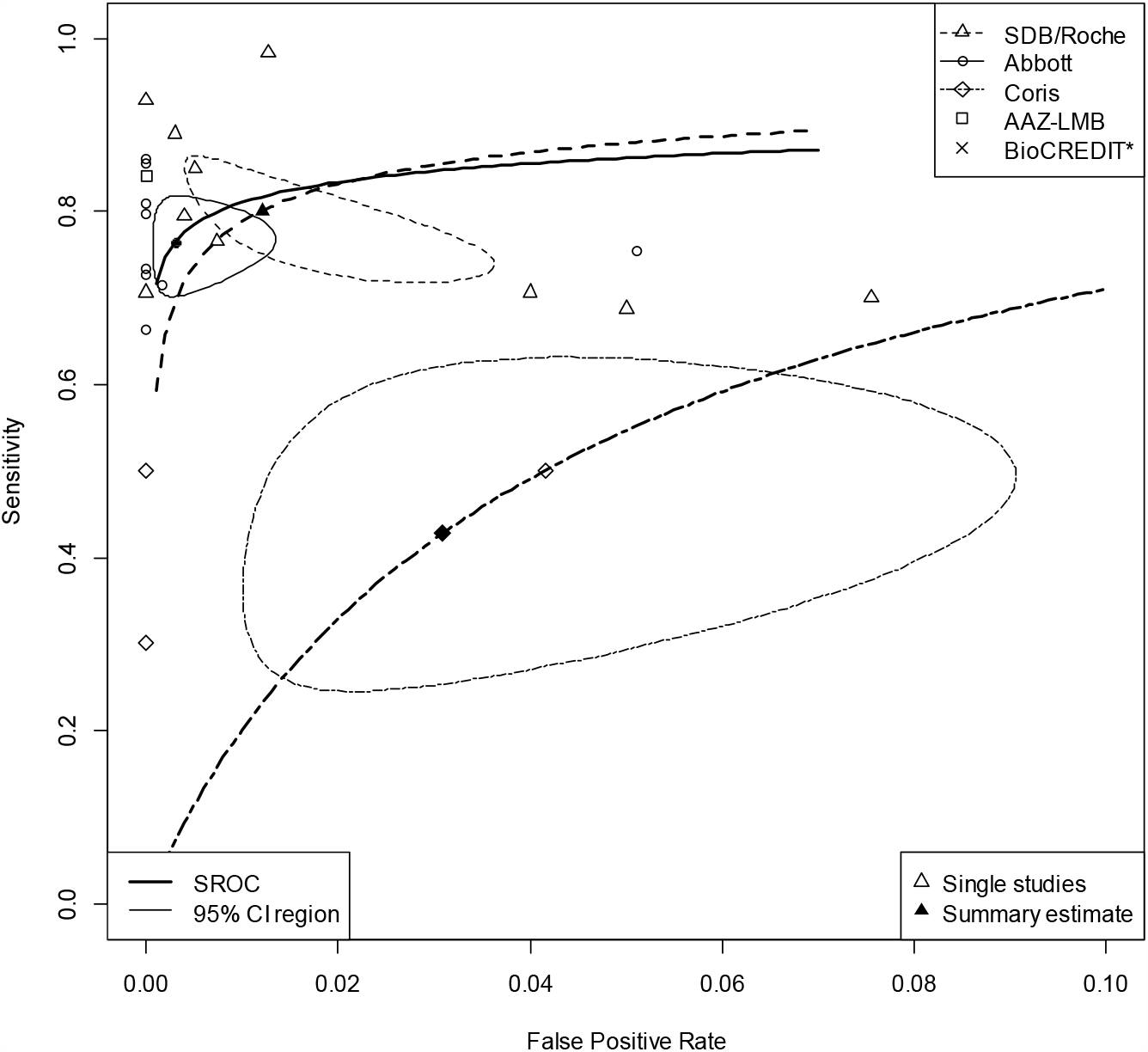
SROC plots for all*. *Study result for BioCREDIT lies outside the plotting region (x=0.55, y=0.525) and is therefore not shown.

**Figure 4.**
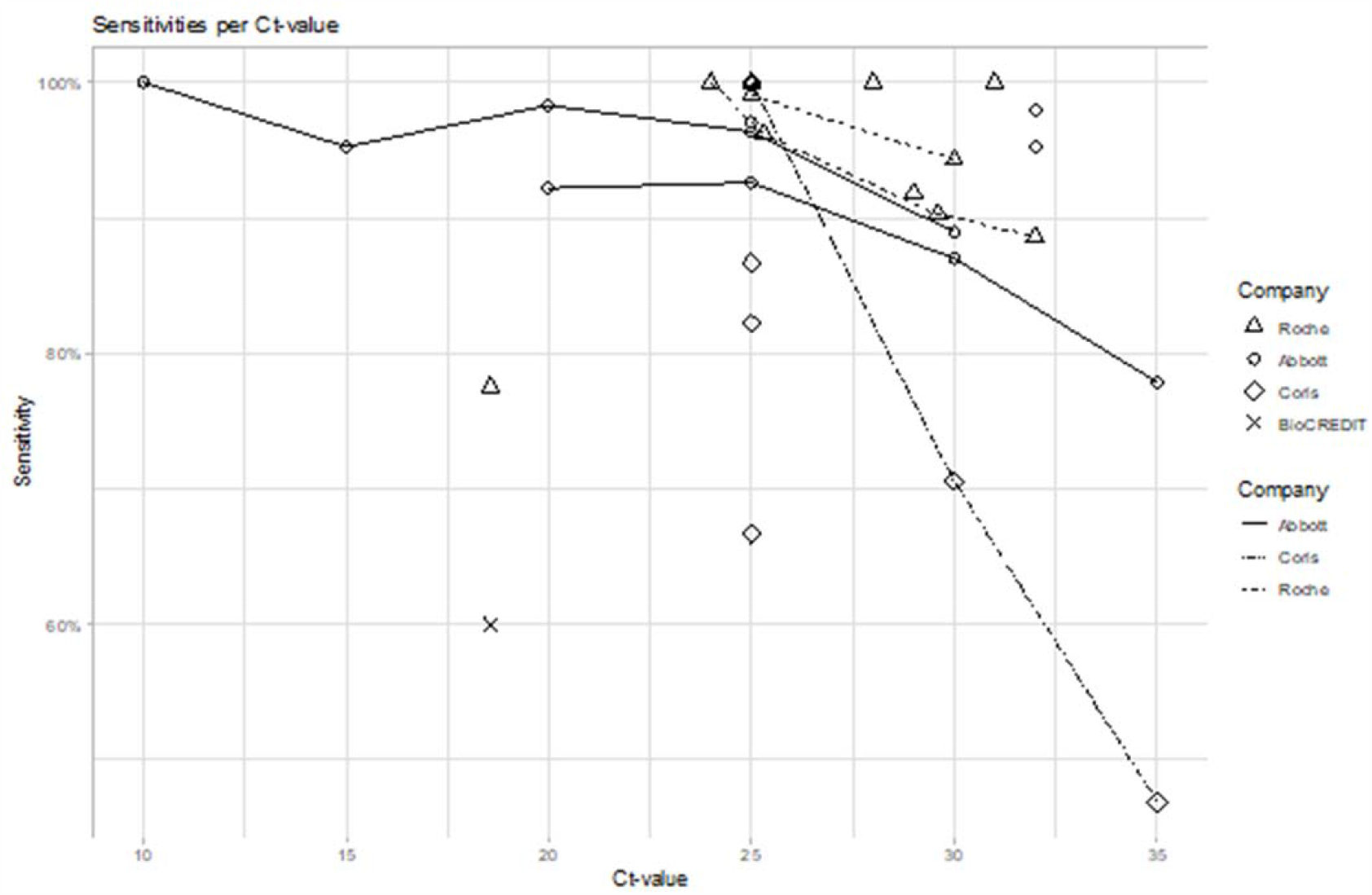
Rapid antigen test sensitivity according to viral load, represented by Ct value groups.

### Critical appraisal of data comparability

Our review presents an overview of available manufacturer-independent, clinical performance data of commercial SARS-CoV-2 RATs not requiring a reading instrument. Altogether, 19 studies investigating five different RATs presented detailed population characteristics and Ct values. A rather comprehensive database exists for two assays, while the other three tests present insufficient data and do not meet the WHO requirement thresholds for sensitivity and specificity.

There is one major concern to be highlighted when comparing performance data of RATs originating from different performance studies:

Data presented in different trials must not directly be compared with each other as numerous variables impact on the resulting performance values. A direct comparison can only be made within one trial, performing a head-to-head comparison of different tests within an identical setting or when using several studies such that effects average out. Several variables impact considerably on clinical performance numbers and prohibit any direct comparison:

1. <u>Preanalytical influencers</u>
  a. Sample type and way of sampling (same or opposite nostril, combined oro-/nasopharyngeal vs nasopharyngeal sample only, order of sampling)
  b. Collection device, transport media and volume vs direct testing without dilution by transport media
  c. Time to test and storage/transport conditions, time delay before processing
2. <u>Analytical influencers</u>
  a. Viral load of the sample and viral load distribution in the respective cohort, represented by Ct or RNA copies/mL
  b. Analytical sensitivity and specificity of the assay
  c. PCR assay specifics, different target genes (E-/RdRp-/N-gene etc.)
  d. Across-laboratory differences (e.g. definition of a positive sample starting at Ct <38 or <40)
3. <u>Clinical parameters of the tested subject</u>
  a. Days post symptom onset of sampling
  b. Asymptomatic vs symptomatic status, definition of symptoms “suspective of SARS-Cov-2 infection”
  c. Severity of symptom presentation

It must be noted that symptom classifications considerably differed between the studies. One study even investigated different populations for self-reported versus physician-defined symptoms. A uniform definition of “clinical symptoms suggestive for SARS-CoV-2 infection” would be desirable but is currently not available.

Due to methodological reasons, the detection limit for SARS-CoV-2 RNA material out of clinical samples tested by RT-PCR is always lower than the detection limit for SARS-Cov-2 antigen. With decreasing viral load, the detectability of even the best performing RAT deteriorates. Cell culture studies on discrepant test results (positive on RT-PCR, negative on RAT) show rare to no virus growth at such samples with high Ct values.^9,28^ This translates into very limited to no infectiousness of the infected patients, even if RT-PCR may still show positive signals for up to three more weeks after peak Ct value.^29,30^

## Conclusion

Only two RATs offer sufficient real-world and manufacturer-independent performance data to recommend their use for the detection of current SARS-CoV-2 infection in symptomatic or suspected high-viral-load patient populations. A high positive predictive value requires testing at a high pre-test probability setting. This requires careful preselection and confirmation of recent contact to confirmed cases and/or knowledge about the underlying local population prevalence. If RATs are used for screening of asymptomatic cases in low-prevalence scenarios, a lower positive predictive value of the according result has to be considered.

## Data Availability

The data supporting this meta-analysis are from previously reported studies and datasets which have been cited. The processed data are available from the corresponding author upon request.

## Acknowledgements

Editorial assistance was provided by Elements Communications Ltd, Westerham, UK and funded by Roche Diagnostics.

